# Clinical features and the maternal and neonatal outcomes of pregnant women with coronavirus disease 2019

**DOI:** 10.1101/2020.03.22.20041061

**Authors:** Rui Nie, Shao-shuai Wang, Qiong Yang, Cui-fang Fan, Yu-ling Liu, Wen-cong He, Mei Jiang, Cheng-cheng Liu, Wan-jiang Zeng, Jian-li Wu, Kutluk Oktay, Ling Feng, Lei Jin

## Abstract

**BACKGROUND:** There is little information about the coronavirus disease 2019 (Covid-19) during pregnancy. This study aimed to determine the clinical features and the maternal and neonatal outcomes of pregnant women with Covid-19.

**METHODS:** In this retrospective analysis from five hospitals, we included pregnant women with Covid-19 from January 1 to February 20, 2020. The primary composite endpoints were admission to an intensive care unit (ICU), the use of mechanical ventilation, or death. Secondary endpoints included the clinical severity of Covid-19, neonatal mortality, admission to neonatal intensive care unit (NICU), and the incidence of acute respiratory distress syndrome (ARDS) of pregnant women and newborns.

**RESULTS:** Thirty-three pregnant women with Covid-19 and 28 newborns were identified. One (3%) pregnant woman needed the use of mechanical ventilation. No pregnant women admitted to the ICU. There were no moralities among pregnant women or newborns. The percentages of pregnant women with mild, moderate, and severe symptoms were 13 (39.4%),19(57.6%), and 1(3%). One (3.6%) newborn developed ARDS and was admitted to the NICU. The rate of perinatal transmission of SARS-CoV-2 was 3.6%.

**CONCLUSIONS:** This report suggests that pregnant women are not at increased risk for severe illness or mortality with Covid-19 compared with the general population. The SARS-CoV-2 infection during pregnancy might not be associated with as adverse obstetrical and neonatal outcomes that are seen with the severe acute respiratory syndrome coronavirus (SARS-CoV) and Middle East respiratory syndrome coronavirus (MERS-CoV) infection during pregnancy. (Funded by the National Key Research and Development Program.)

## INTRODUCTION

The outbreak of the coronavirus disease 2019 (Covid-19) has spread quickly from Wuhan, China to all other major cities in China and is rapidly spreading to a growing number of other countries despite efforts at containment and isolation^1^. The population is generally susceptible to this newly identified coronavirus^2^, initially termed 2019-nCoV and subsequently SARS-CoV-2. Early efforts have focused on describing the clinical characteristics and outcomes of Covid-19 in the general population^1,3,4^. However, the disease appears to be particularly impactful in special populations that include those older than sixty-five years of age ^5^. Pregnant women are also considered to be a special population group because of the unique ‘immune suppression’ caused by pregnancy^6^. The immunologic and physiologic changes might make pregnant women at higher risk of severe illness or mortality with Covid-19, compared with the general public. However, there is little information on Covid-19 during pregnancy. This study aimed to determine the clinical features and the maternal and neonatal outcomes of pregnant women with Covid-19.

## METHODS

### STUDY DESIGN AND PARTICIPANT

This is a retrospective analysis of data from multiple centers. From January 1 to February 20, 2020, all consecutive pregnant women with Covid-19 in five hospitals were enrolled. The five hospitals were Tongji Hospital of Huazhong University of Science and Technology, Renmin Hospital of Wuhan University, Maternal and Child Health Hospital of Hubei Province, Yichang Central People’s Hospital, and Jinmen Second People’s Hospital. Among the five hospitals, three hospitals were in Wuhan, and two hospitals were in other cities of the Hubei Province that are adjacent to Wuhan and in endemic areas of Covid-19. All five hospitals assigned by the government to be responsible for the treatments of pregnant women with Covid-19. The Institutional Review Board of Huazhong University of Science and Technology approved this study (#TJ-IRB20200222). All pregnant women gave informed consent to participate in this study.

### LABORATORY CONFIRMATION

Based on the New Coronavirus Pneumonia Prevention and Control Program (6th edition) published by the National Health Commission of China^7^, those with one of the following laboratory evidence is considered to have a confirmed Covid-19 case: (1) positive for SARS-CoV-2 nucleic acid by real-time reverse-transcription–polymerase-chain-reaction (RT-PCR); (2) viral gene sequencing showing highly homogeneity to the known SARS-CoV-2. Real-time RT-PCR assays were performed following the protocol established by the WHO^8^.

### DATA COLLECTION

We collected data of all pregnant women and newborns from electronic medical records with data collection forms. If there were deficiencies in the data, we directly communicated with the pregnant women and/or their family members by telephone to acquire additional information. The information collected included demographic data, exposure history, past medical history, clinical presentation, laboratory findings, chest computed tomographic (CT) scans, and treatment received.

According to the New Coronavirus Pneumonia Prevention and Control Program (6th edition) published by the National Health Commission of China^7^, the clinical severity of Covid-19 is classified into four levels: (1) Mild: Presence of mild clinical symptoms without CT/X-ray abnormalities. (2) Moderate: Presence of fever or upper respiratory symptoms, and CT/X-ray imaging consistent with pulmonary infection. (3) Severe: Presence of one of three conditions: i) shortness of birth, and respiratory rate≥30 breaths per minute; ii) resting Oxygen Saturation≤93%; iii) partial pressure of oxygen/fraction of inspired oxygen ≤ 300mmHg. (4) Severe acute: Presence of acute respiratory distress and need for mechanical ventilation, or presence of shock, or admission to the intensive care unit (ICU) because of the development of organ dysfunction.

Exposure history is defined as a history of travel to or residence in Wuhan or other cities with continuous transmission of local cases in the last 14 days before the onset of symptoms, contact with patients with fever or respiratory symptoms from Wuhan or other cities with continuous transmission of local cases in the last 14 days before the onset of symptom, or being epidemiologically connected to SARS-CoV-2 infections or clustered onsets^2^.

The Pregnancy outcome was classified based on the gestational age at delivery (<37 weeks or ≥37 weeks), mode of delivery (vaginal or cesarean section), indication for cesarean section, and obstetrical complications. Information collected from newborns included birth weight, Apgar score, neonatal death, tests for SARS-CoV-2, and manifestations of Covid-19. Low birth weight was defined as a birth weight of less than 2,500 grams^9^. Very low birth weight was defined as a birth weight of less than 1,500 grams^9^.

### STUDY OUTCOMES

The primary composite endpoints were admission to an ICU, the use of mechanical ventilation, or death. Secondary endpoints included the clinical severity of Covid-19, neonatal mortality, admission to neonatal intensive care unit (NICU), and the incidence of acute respiratory distress syndrome (ARDS) of pregnant women or newborns.

### STATISTICAL ANALYSIS

Patient characteristics were presented using descriptive statistics. Continuous variables were summarized using the mean and standard deviations, while categorical variables were calculated using the frequency and percentage. All statistical analyses were performed using SAS 9.4 (SAS Institute, Cary, NC).

## RESULTS

### CHARACTERISTICS OF PREGNANT WOMEN

Our retrospective analysis revealed a total of 33 pregnant women with Covid-19 who were admitted to 5 public hospitals in Hubei Province. The maternal age ranged from 24 to 36 years. Among the affected, there were 8 (24.2%) health care workers, and 32 (97%) of women had a history of exposure. Three women were in the second trimester (17 weeks, 20 weeks, and 26 weeks), and the other 30 women were in the third trimester at the time of presentation. Fifteen (45.5%) women had chronic disorders involving cardiovascular and cerebrovascular, digestive, endocrine, and nervous systems, as well as infectious diseases and depressive disorders (Table 1).

**Table 1.**
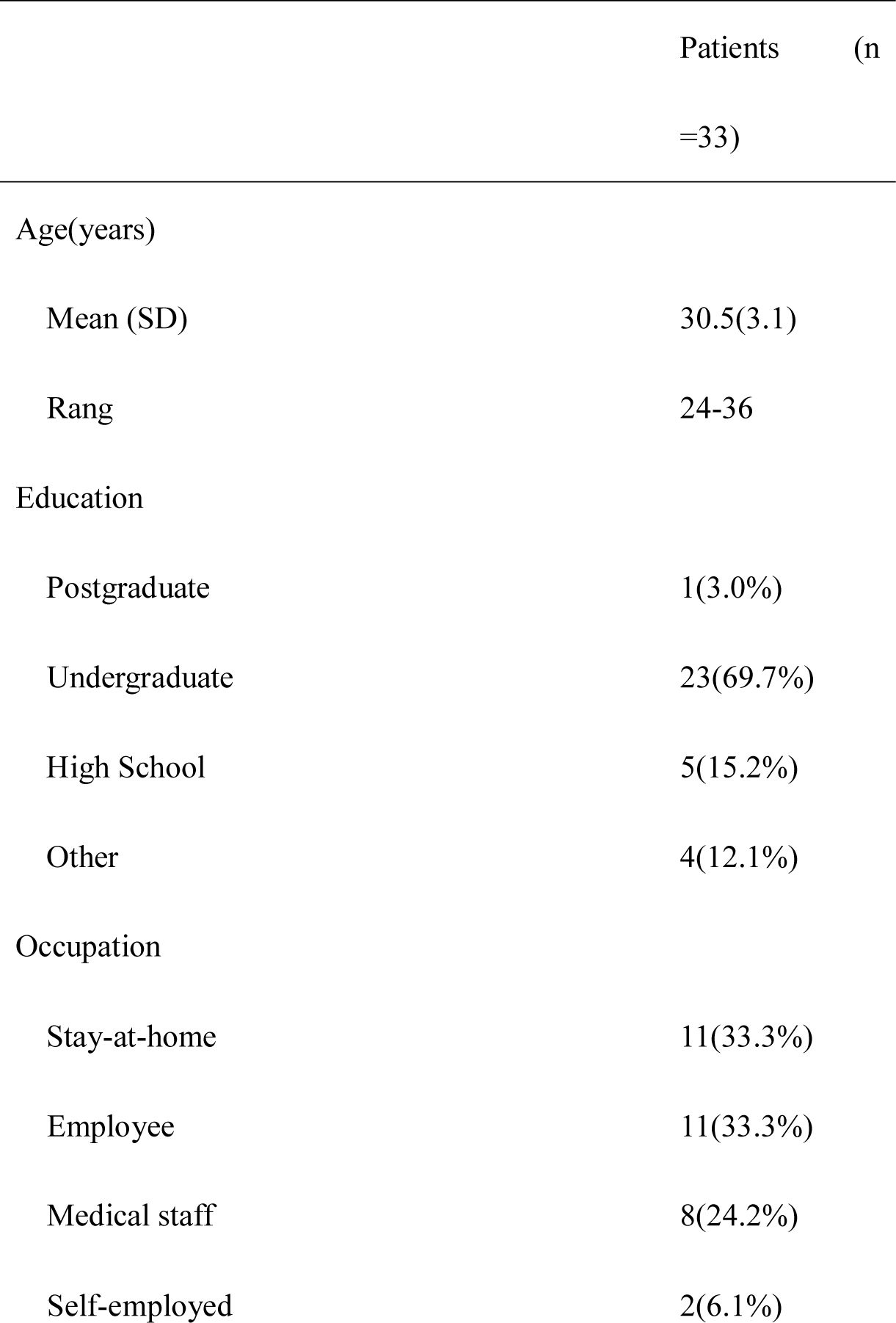
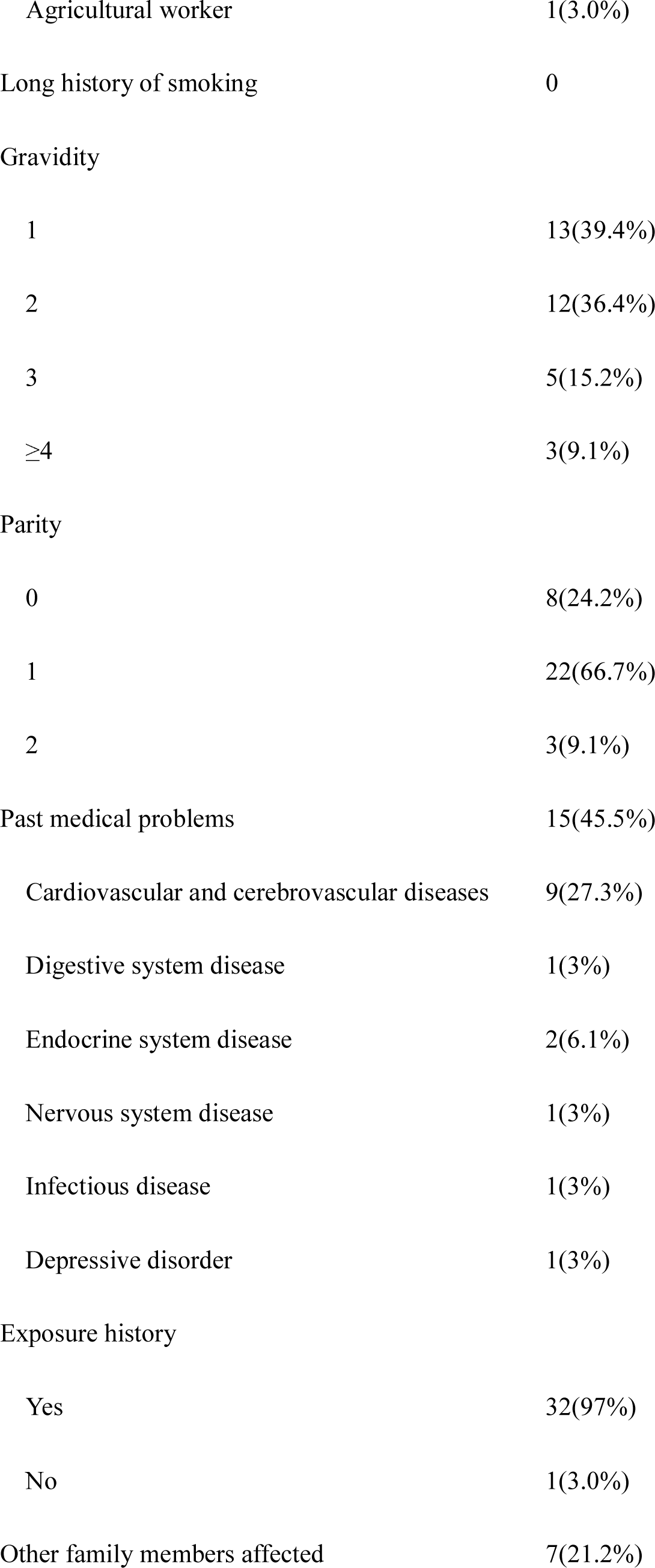
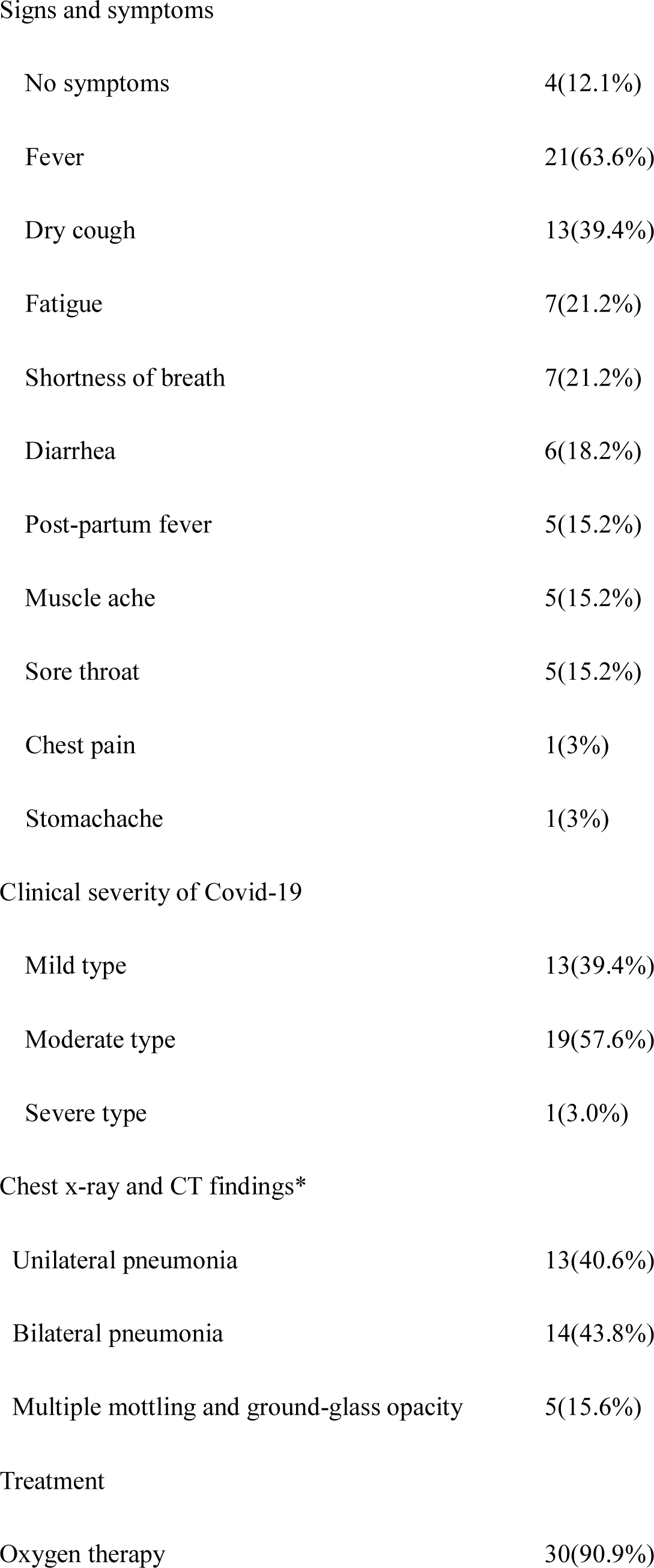
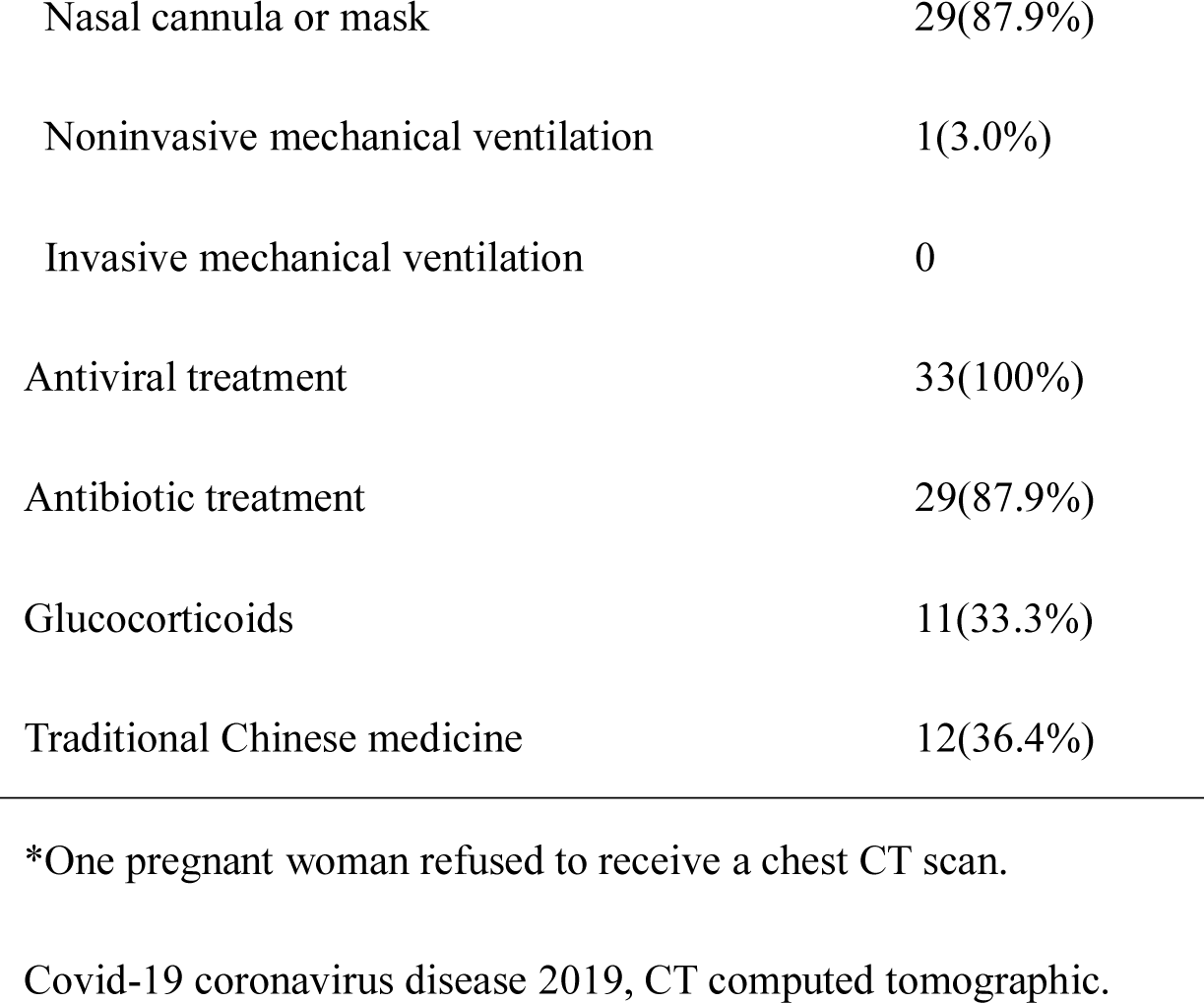
Characteristics, clinical presentation, and treatment received for pregnant women with Covid-19

|  | Patients (n=33) |
| --- | --- |
| Age(years) |  |
| Mean (SD) | 30.5(3.1) |
| Rang | 24-36 |
| Education |  |
| Postgraduate | 1(3.0%) |
| Undergraduate | 23(69.7%) |
| High School | 5(15.2%) |
| Other | 4(12.1%) |
| Occupation |  |
| Stay-at-home | 11(33.3%) |
| Employee | 11(33.3%) |
| Medical staff | 8(24.2%) |
| Self-employed | 2(6.1%) |
| Agricultural worker | 1(3.0%) |
| Long history of smoking | 0 |
| Gravidity |  |
| 1 | 13(39.4%) |
| 2 | 12(36.4%) |
| 3 | 5(15.2%) |
| ≥4 | 3(9.1%) |
| Parity |  |
| 0 | 8(24.2%) |
| 1 | 22(66.7%) |
| 2 | 3(9.1%) |
| Past medical problems | 15(45.5%) |
| Cardiovascular and cerebrovascular diseases | 9(27.3%) |
| Digestive system disease | 1(3%) |
| Endocrine system disease | 2(6.1%) |
| Nervous system disease | 1(3%) |
| Infectious disease | 1(3%) |
| Depressive disorder | 1(3%) |
| Exposure history |  |
| Yes | 32(97%) |
| No | 1(3.0%) |
| Other family members affected | 7(21.2%) |
| Signs and symptoms |  |
| No symptoms | 4(12.1%) |
| Fever | 21(63.6%) |
| Dry cough | 13(39.4%) |
| Fatigue | 7(21.2%) |
| Shortness of breath | 7(21.2%) |
| Diarrhea | 6(18.2%) |
| Post-partum fever | 5(15.2%) |
| Muscle ache | 5(15.2%) |
| Sore throat | 5(15.2%) |
| Chest pain | 1(3%) |
| Stomachache | 1(3%) |
| Clinical severity of Covid-19 |  |
| Mild type | 13(39.4%) |
| Moderate type | 19(57.6%) |
| Severe type | 1(3.0%) |
| Chest x-ray and CT findings\* |  |
| Unilateral pneumonia | 13(40.6%) |
| Bilateral pneumonia | 14(43.8%) |
| Multiple mottling and ground-glass opacity | 5(15.6%) |
| Treatment |  |
| Oxygen therapy | 30(90.9%) |
| Nasal cannula or mask | 29(87.9%) |
| Noninvasive mechanical ventilation | 1(3.0%) |
| Invasive mechanical ventilation | 0 |
| Antiviral treatment | 33(100%) |
| Antibiotic treatment | 29(87.9%) |
| Glucocorticoids | 11(33.3%) |
| Traditional Chinese medicine | 12(36.4%) |

\*One pregnant woman refused to receive a chest CT scan.
Covid-19 coronavirus disease 2019, CT computed tomographic.

Among the 33 pregnant women, the most common symptoms were fever (21[63.6%]), dry cough (13 [39.4%]), fatigue (7 [21.2%]), and shortness of breath (7 [21.2%]). Less common symptoms were diarrhea, post-partum fever, muscle ache, sore throat, chest pain, and stomachache. Four (12.1%) pregnant women had no obvious symptoms. The clinical severity of most women was mild (13[39.4%]) and moderate (19[57.6%]). Only one (3%) woman proceeded to a severe type of COVID-19 (Table 1).

By chest CT, 13 (40.6%) women showed bilateral pneumonia, 14 (43.8%) had showed unilateral pneumonia, and 5 (15.6%) showed multiple mottling and ground-glass opacity (Table 1).

Twenty-nine (87.9%) pregnant women required oxygen supplementation via a nasal cannula or mask, and one (3%) patient needed noninvasive mechanical ventilation. All women received antiviral therapy. Twenty-nine (87.9%) women were given antibiotic treatment, 11 (33.3%) were treated with glucocorticoids, and 12 (36.4%) received traditional Chinese medicine (Table 1).

### MATERNAL OUTCOMES

Twenty-two (81.5%) women delivered via cesarean section, and 5(18.5%) had vaginal deliveries. The overall rate of obstetrical complications was 22.2%, including three cases of preterm premature rupture of membranes, two cases of hypertensive diseases of the pregnancy, two cases of gestational diabetes mellitus, and one case of spontaneous preterm labor. As February 20, 2020, five pregnancies were ongoing, and one woman decided to terminate her pregnancy for personal reasons (Table 2).

**Table 2.** Maternal outcomes

|  | Patients (n =33) |
| --- | --- |
| Maternal outcome |  |
| ICU admission | 0 |
| Death | 0 |
| Remained in hospital | 24(72.7%) |
| Discharged | 9(27.3%) |
| Mode of delivery |  |
| Vaginal delivery | 5(18.5%) |
| Cesarean section | 22(81.5%) |
| Ongoing pregnancy | 5(15.2%) |
| Induced abortion | 1(3.0%) |
| Obstetrical complications in general | 6(22.2%) |
| Preterm premature rupture of membranes | 3(11.1%) |
| hypertensive diseases of the pregnancy | 2(7.4%) |
| Gestational diabetes mellitus | 2(7.4%) |
| Spontaneous preterm labor | 1(3.7%) |

ICU intensive care unit.

By February 20, 2020, 9 (27.3%) women had been discharged while the remaining were still hospitalized. No pregnant women proceeded to severe complications or were admitted to the ICU, and there was no mortality.

### NEONATAL OUTCOMES

There were 28 newborns born to 27 women, ten (35.7%) of whom were delivered preterm. The 1-min Apgar score of all newborns ranged from 8-10. The 5-min Apgar score of all newborns ranged from 9-10. All newborns were isolated after birth and fed by milk formula. Five (17.9%) newborns had low birth weight, and no newborn had very low birth weight. Four (14.3%) newborns were diagnosed with fetal distress. One newborn who was born at 34 weeks of gestation, was transferred to the NICU with a diagnosis of ARDS. At the end of the follow-up, this newborn recovered and was discharged. No newborns had clinical symptoms of Covid-19 or died (Table 3).

**Table 3.**
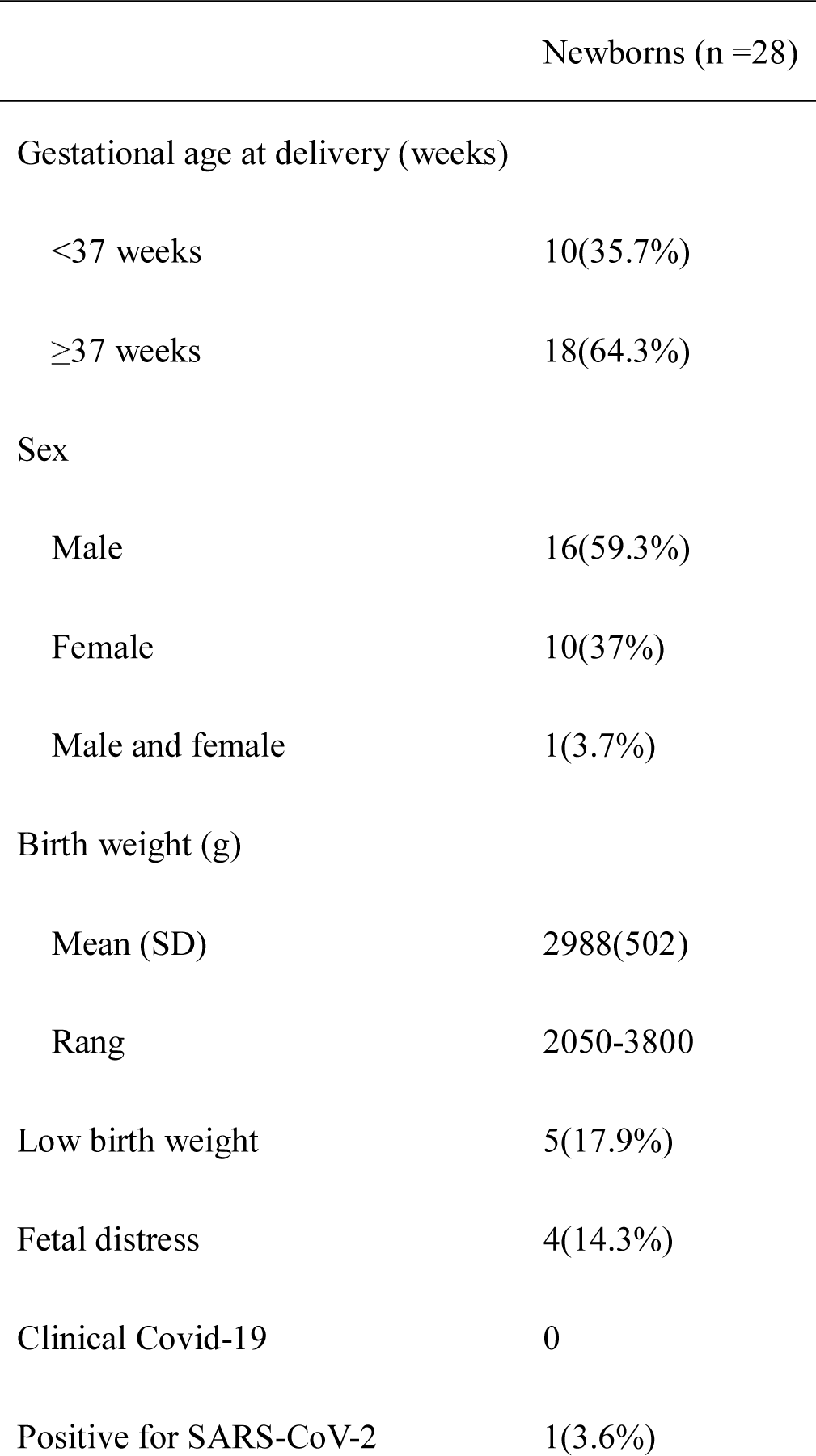
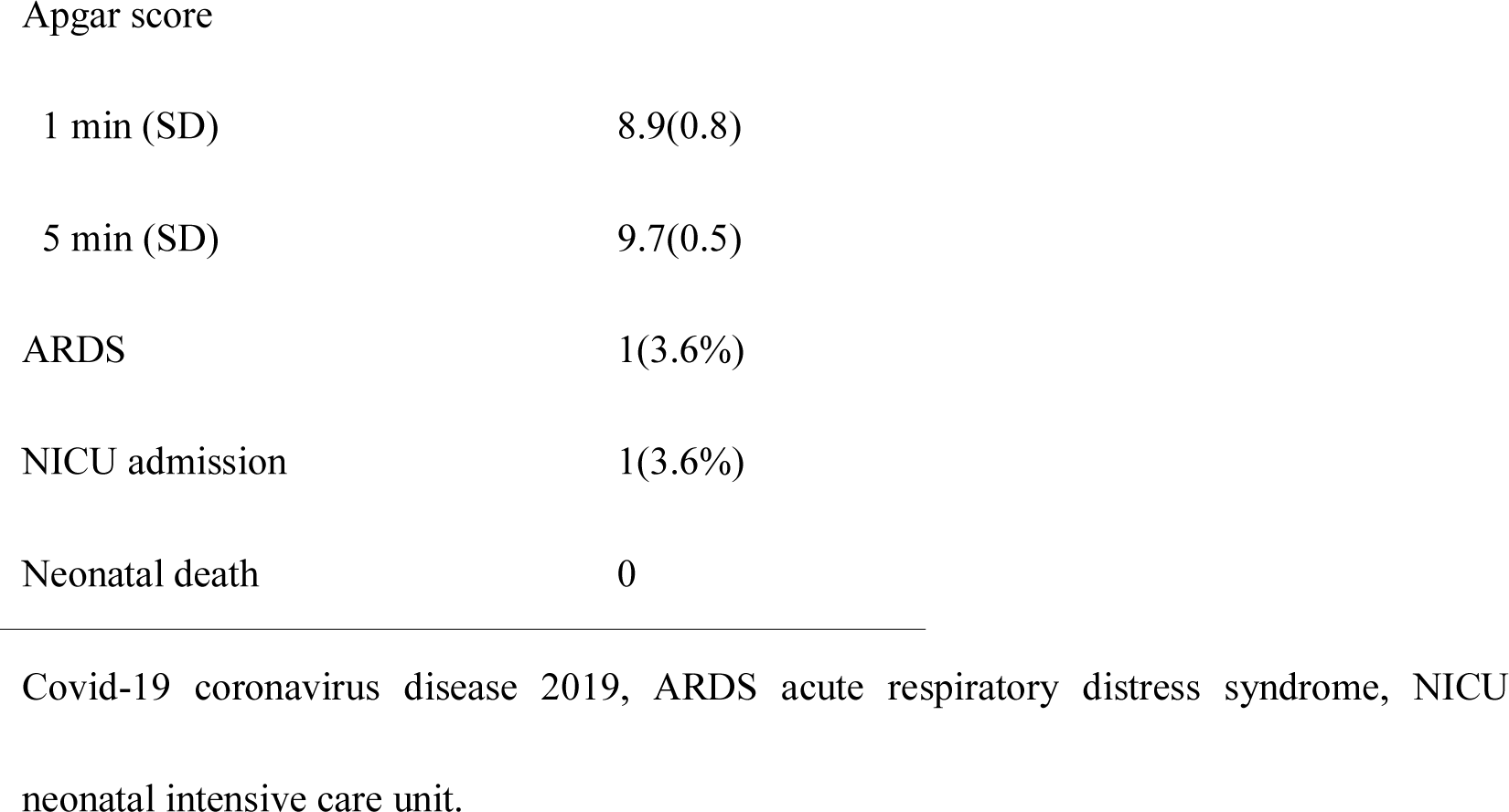
Neonatal outcomes

At the end of the follow-up, two newborns had not been tested for SARS-CoV-2 because their parents asked to delay the test to 14 days after birth. Twenty-six newborns were tested for SARS-CoV-2 by real-time RT-PCR assays of throat swabs. Among those, one newborn tested positive for SARS-CoV-2 infection, thus yielding a perinatal transmission rate of 3.6%. The mother of this newborn was sent to the hospital because of a fever. The mother wore N95 masks during the cesarean section. All medical staff wore protective suits. The newborn had no contact with his mother and other family members. He was directly transferred to the neonatal department and isolated immediately after birth. A throat swab from this mother was collected for the nucleic acid test when she was admitted to the hospital. However, the result was not reported until 36 h after birth because of the massive number of samples that needed testing during the Covid-19 outbreak. Real-time RT-PCR showed that the mother was positive for SARS-CoV-2 infection. At the same time, the throat swab sample was taken from the newborn for a nucleic acid test, which also came back positive. In addition, the chest X-ray of the newborn 53 h after birth was consistent with pulmonary infection. However, the cord blood and placental samples were both negative for SARS-CoV-2 infection. The newborn had no clinical manifestations of Covid-19. The newborn was retested and found to negative for SARS-CoV-2 infection on days 4, 8, and 15 after birth as detected by the nucleic acid test of the real-time RT-PCR of subsequent throat swabs. The chest X-ray of the newborn showed that pulmonary infection had resolved. The newborn did not receive any treatment and was discharged 16 days after birth.

## DISCUSSION

In this study, we reported a case series of pregnant women with Covid-19 and newborns born to these women. The primary composite endpoint occurred in one (3%) pregnant woman who needed the use of mechanical ventilation, while no pregnant women admitted to the ICU or died. For the secondary outcomes, the percentages of pregnant women with mild, moderate, and severe symptoms were 13 (39.4%),19(57.6%), and 1(3%). One (3.6%) newborn developed ARDS and was admitted to the NICU. There were no moralities among newborns.

There is limited knowledge regarding Covid-19 that occurs during pregnancy. What is known has been bases on the experiences from two different coronaviruses, severe acute respiratory syndrome coronavirus (SARS-CoV) and Middle East respiratory syndrome coronavirus (MERS-CoV)^6,10,11^. The clinical outcomes among pregnant women with SARS or MERS were worse than those occurring in non-pregnant women^6,10^. However, our data suggest the contrary to Covid-19. Among the 33 pregnant women with confirmed Covid-19, 39.4% of women presented mild, 57.6% presented moderate, and 3% presented with severe symptoms. For most pregnant women (87.9%), oxygen therapy supplied via a nasal cannula or mask was sufficient for supportive care. Only one (3%) pregnant woman needed the use of mechanical ventilation. At the end of the follow-up, no pregnant women had severe complications or needed to be transferred to the ICU or died. Compared with the general population, in which 5.0% who were admitted to the ICU, 2.3% who needed mechanical ventilation, and 1.4% who died^12^, it seems that the severity and mortality rate of Covid-19 in pregnant women are lower.

The risk of perinatal transmission of SARS-CoV-2 was low, as only one out of 26 newborns was confirmed to have a SARS-CoV-2 infection. In our study, this newborn’s SARS-CoV-2 infection was confirmed by the detection of virus in throat swab and chest X-ray abnormalities. However, we cannot prove that this is a case of vertical transmission because the cord blood and placental samples were both negative for SARS-CoV-2.

How SARS-CoV-2 invades the human body remains a mystery. Xu *et al*. first reported that SARS-CoV-2 shares 76.47% amino acid sequence identity with SARS-CoV, and may use the same receptor, Angiotensin-converting enzyme 2 (ACE2), for entry into target host cells^13^. Zhao *et al*. found that ACE2 is specifically expressed in type II alveolar epithelial cells (AT2) in human lung, suggesting that SARS-CoV-2 may target ACE2-positive AT2 cells to induce pneumonia^14^. Further studies showed that ACE2 protein is abundantly present in humans in the epithelia of the lung and small intestine^15^, heart^16^, arterial smooth muscle cells, gastrointestinal system, renal tubules of kidney^17^ and testes ^18^, suggesting potential routes for SARS-CoV-2 infection in the respiratory, cardiovascular, gastrointestinal, urinary and male reproductive systems. Thus, this newborn could have been infected from many possible paths, such as via the respiratory system.

However, this newborn’s case may also be real vertical transmission of SARS-CoV-2 since the mother wore N95 masks during the cesarean section, and all medical staff wore protective suits. Furthermore, the newborn was isolated immediately after birth. These preventive measures should have decreased the risk of SARS-CoV-2 transmission through the respiratory system of the mother, medical staff, and hospital environment. Second, previous studies have shown that ACE2 is expressed by the human endometrium and placenta ^19,20^. A recent study revealed that ACE2 is highly expressed in maternal-fetal interface cells, including stromal cells and perivascular cells of decidua, and cytotrophoblast and syncytiotrophoblast in placenta^21^. Thus, SARS-CoV-2 may also be attracted to the human endometrium and placenta though ACE2, causing vertical transmission of SARS-CoV-2. Also, the SARS-CoV-2 has been detected in urine and stool^22^, suggesting a possibility of ascending infection of the uterus, placenta, and fetus. Third, it has been reported that the ACE2-expressing cell ratio is variable between individuals, which may explain why some people are more vulnerable to SARS-CoV-2^14^. This characteristic of ACE2 may explain why SARS-CoV-2 infected only one out of 26 newborns.

Another novelty of our study, the clinical characteristics and outcomes of infected newborns have never been reported. This 36-hour-old newborn is the youngest baby ever to be reported with SARS-CoV-2 infection. Interestingly, the newborn with SARS-CoV-2 infection did not have any clinical symptoms. He converted to seronegative in 2 days and recovered without treatment. This limited information suggests that the newborns may generally be resistant to infection with SAR-Cov-2 and, if infected, likely not suffer from significant consequences.

During the SARS outbreak, Wong *et al*. evaluated the obstetrical outcomes from a cohort of pregnant women with SARS in Hong Kong^6^. Four of the seven patients who presented in the first trimester had a spontaneous miscarriage. Four of the five patients who presented after 24 weeks delivered preterm. Two mothers recovered without delivery, but their ongoing pregnancies were complicated by intrauterine growth restriction^6^. In our study, we included three women in the second trimester and 30 women in the third trimester at presentation. Among the 33 women, 27 women delivered 28 newborns, and the live birth rate was 100%. Five women had ongoing pregnancy without severe obstetrical complications at the time of manuscript preparation. The overall rate of obstetrical complications was 22.2%, including three cases of preterm premature rupture of membranes, two cases of hypertensive diseases of the pregnancy, two cases of gestational diabetes mellitus, and one case of spontaneous preterm labor. Therefore, the obstetrical outcomes from pregnant women with SARS-CoV-2 infection appear better than that for pregnant women with SARS. However, it’s worth noting that most pregnant women were infected with SARS-CoV-2 during the third trimester in this study. The gestational age when women are infected may affect obstetrical outcomes.

In a review of 11 pregnant women with MERS-CoV infection, the infant death rate was 27%^11^. For SARS during pregnancy, the fetal/neonatal complications included preterm labor, intrauterine growth restriction, intrauterine demise, and neonatal death ^6^. In this current study, no newborn infants died. The preterm birth rate was 35.7%, and the rate of low birth weight was 17.9%. Four (14.3%) of 28 newborn infants had signs of fetal distress. One newborn infant who was born at 34 weeks developed ARDS and was admitted to the NICU. At the end of the follow-up, this newborn infant had recovered and was discharged. Our experience showed that SARS-CoV-2 infection during pregnancy might not be associated with as adverse neonatal outcomes as SARS-CoV and MERS-CoV infection during pregnancy.

The clinical features of Covid-19 in pregnant women has not yet been reported. In this study, we report some clinical features of pregnant women with Covid-19: First, four (12.1%) pregnant women had no apparent symptoms during hospitalization. However, they were positive for SARS-CoV-2 by the real-time RT-PCR assay. This finding suggests that there are some asymptomatic carriers of SARS-CoV-2 during pregnancy. Because asymptomatic carriers of SARS-CoV-2 may transmit virus^23^, we recommend that all pregnant women should be tested for SARS-CoV-2 during the Covid-19 outbreak. Second, in our study, five women delivered babies by vaginal birth, and their newborns were negative for SARS-CoV-2. This finding implies that vaginal delivery may not increase the risk of SARS-CoV-2 transmission to newborns. Therefore, Covid-19 should not be the sole indication for the cesarean section. The timing and mode of delivery should depend on the gestational age, the maternal and fetal condition.

This observational study consisted of 33 pregnant women and 28 newborns. Although this report is the most extensive case series so far, the sample size is small. We caution our readers to take this into account when interpreting the results and conclusions drawn from this study, since some of the above observations may be due to chance occurrence. However, there is currently no large series of pregnant women with confirmed Covid-19, so we believe that this study will be useful for guiding us in the management of pregnant women with Covid-19 and their newborns.

In conclusion, pregnant women are not at increased risk for severe illness or mortality with Covid-19 compared with the general population. The SARS-CoV-2 infection during pregnancy might not be associated with as adverse obstetrical and neonatal outcomes that are seen with the SARS-CoV and MERS-CoV infection during pregnancy. There appears to be a low risk of perinatal transmission of SARS-CoV-2. However, we should not rule out the potential risk of vertical transmission of SARS-COV-2.

## Data Availability

The data that support the findings of this study are available from the corresponding author, upon reasonable request.

Supported by the National Key Research and Development Program (number 2018YFC1002103).

All authors declare no competing interests.

We thank all people of combatting Covid-19.

## REFERENCES

1. Huang C, Wang Y, Li X, et al. Clinical features of patients infected with 2019 novel coronavirus in Wuhan, China. Lancet 2020; 395:497–506.

2. Jin Y-H, Cai L, Cheng Z-S, et al. A rapid advice guideline for the diagnosis and treatment of 2019 novel coronavirus (2019-nCoV) infected pneumonia (standard version). Mil Med Res 2020; 7: 4.

3. Chen N, Zhou M, Dong X, et al. Epidemiological and clinical characteristics of 99 cases of 2019 novel coronavirus pneumonia in Wuhan, China: a descriptive study. Lancet 2020;395: 507–13.

4. Wang D, Hu B, Hu C, et al. Clinical Characteristics of 138 Hospitalized Patients With 2019 Novel Coronavirus-Infected Pneumonia in Wuhan, China. JAMA 2020; Published online February 7. Doi:10.1001/jama.2020.1585.

5. Yang X, Yu Y, Xu J, et al. Clinical course and outcomes of critically ill patients with SARS-CoV-2 pneumonia in Wuhan, China: a single-centered, retrospective, observational study. Lancet Respir Med 2020. Published online Feb 24. Doi: 10.1016/S2213-2600(20)30079-5.

6. Wong SF, Chow KM, Leung TN, et al. Pregnancy and perinatal outcomes of women with severe acute respiratory syndrome. Am J Obstet Gynecol 2004; 191: 292–7.

7. National Health Commission of China. New Coronavirus Pneumonia Prevention and Control Program (6th edition). (http://www.nhc.gov.cn/yzygj/s7653p/202002/81BQZKqdp2CV3QV5nUEsqSg1ygegLmqRygj/fifil/b21BQZKqdp2CV3QV5nUEsqSg1ygegLmqRygjb817.pdf).

8. World Health Organization. Coronavirus disease (COVID-19) technical guidance: laboratory testing for 2019-nCoV in humans (https://www.who.int/emergencies/diseases/novel-coronavirus-2019/technical-guidance/laboratory-guidance).

9. Maheshwari A, Pandey S, Shetty A, Hamilton M, Bhattacharya S. Obstetric and perinatal outcomes in singleton pregnancies resulting from the transfer of frozen thawed versus fresh embryos generated through in vitro fertilization treatment: A systematic review and meta-analysis. Fertil Steril 2012; 98: 368–77.

10. Favre G, Pomar L, Musso D, Baud D. 2019-nCoV epidemic: what about pregnancies? Lancet 2020; 395: e40.

11. Alfaraj SH, Al-Tawfiq JA, Memish ZA. Middle East Respiratory Syndrome Coronavirus (MERS-CoV) infection during pregnancy: Report of two cases & review of the literature. J Microbiol Immunol Infect 2019; 52: 501–3.

12. Guan WJ, Ni ZY, Hu Y, et al. Clinical Characteristics of Coronavirus Disease 2019 in China. N Engl J Med 2020; Published online Feb 28. Doi: 10.1056/NEJMoa2002032.

13. Xu X, Chen P, Wang J, et al. Evolution of the novel coronavirus from the ongoing Wuhan outbreak and modeling of its spike protein for risk of human transmission. Sci China Life Sci 2020; published online Jan 26. Doi: 10.1007/s11427-020-1637-5.

14. Zhao Y, Zhao Z, Wang Y, Zhou Y, Ma Y, Zuo W. Single-cell RNA expression profiling of ACE2, the putative receptor of Wuhan 2019-nCov. bioRxiv 2020; published online Jan 21. DOI: https://doi.org/10.1101/2020.01.26.919985.

15. Hamming I, Timens W, Bulthuis MLC, Lely AT, Navis GJ, van Goor H. Tissue distribution of ACE2 protein, the functional receptor for SARS coronavirus. A first step in understanding SARS pathogenesis. J Pathol 2004; 203: 631–7.

16. Crackower MA, Sarao R, Oliveira-dos-Santos AJ, da Costa J, Zhang L. Angiotensin-converting enzyme 2 is an essential regulator of heart function. Nature 2002; 417: 822–8.

17. Fan C, Li K, Ding Y, Lu W, Wang J. ACE2 Expression in Kidney and Testis May Cause Kidney and Testis Damage After 2019-nCoV Infection. medRxiv 2020; published online Feb 13. DOI: https://doi.org/10.1101/2020.02.12.20022418.

18. Zou X, Chen K, Zou J, et al. The single-cell RNA-seq data analysis on the receptor ACE2 expression reveals the potential risk of different human organs vulnerable to Wuhan 2019-nCoV infection. Front Med 2020; published online Feb 8. Doi: 10.1007/s11684-020-0754-0.

19. Wang Y, Lumbers ER, Arthurs AL, et al. Regulation of the human placental (pro)renin receptor-prorenin-angiotensin system by microRNAs. Mol Hum Reprod 2018; 24: 453–64.

20. Vaz-Silva J, Carneiro MM, Ferreira MC, et al. The vasoactive peptide angiotensin-(1-7), its receptor Mas and the angiotensin-converting enzyme type 2 are expressed in the human endometrium. Reprod Sci 2009; 16: 247–56.

21. Li M, Chen L, Xiong C, Li X. The ACE2 expression of maternal-fetal interface and fetal organs indicates potential risk of vertical transmission of SARS-COV-2. bioRxiv 2020; published online Feb 27. DOI: https://doi.org/10.1101/2020.02.27.967760

22. Pan Y, Zhang D, Yang P, Poon LLM, Wang Q. Correspondence Viral load of SARS-CoV-2 in clinical samples. Lancet Infect Dis 2020; published online Feb 24. Doi: 10.1016/S1473-3099(20)30113-4

23. December S. Asymptomatic cases in a family cluster with SARS-CoV-2 infection. Lancet Infect Dis 2020; published online Feb 19. Doi: 10.1016/S1473-3099(20)30114-6.

